# Differential COVID-19-attributable mortality and BCG vaccine use in countries

**DOI:** 10.1101/2020.04.01.20049478

**Authors:** Anita Shet, Debashree Ray, Neelika Malavige, Mathuram Santosham, Naor Bar-Zeev

## Abstract

While mortality attributable to COVID-19 has devastated global health systems and economies, striking regional differences have been observed. The *Bacille Calmette Guérin* (BCG) vaccine has previously been shown to have non-specific protective effects on infections, as well as long-term efficacy against tuberculosis. Using publicly available data we built a simple log-linear regression model to assess the association of BCG use and COVID-19-attributable mortality per 1 million population after adjusting for confounders including country economic status (GDP per capita), and proportion of elderly among the population. The timing of country entry into the pandemic epidemiological trajectory was aligned by plotting time since the 100^th^ reported case. Countries with economies classified as lower-middle-income, upper-middle-income and high-income countries (LMIC, UMIC, HIC) had median crude COVID-19 log-mortality of 0.4 (Interquartile Range (IQR) 0.1, 0.4), 0.7 (IQR 0.2, 2.2) and 5.5 (IQR 1.6, 13.9), respectively. COVID-19-attributable mortality among BCG-using countries was 5.8 times lower [95% CI 1.8-19.0] than in non BCG-using countries. Notwithstanding limitations due to testing constraints in LMICs, case ascertainment bias and a plausible rise of cases as countries progress along the epidemiological trajectory, these analyses provide intriguing observations that urgently warrant mobilization of resources for prospective randomized interventional studies and institution of systematic disease surveillance, particularly in LMICs.

## Introduction

Novel SARS-CoV2 continues to wreak global havoc. Mortality is of greatest concern directly influencing national response and policy. Early reports from Hubei province in China reported a case fatality rate (CFR) of 15%^1^, which with widening surveillance, rapidly decreased to below 3%^2,3^. As new epidemics began in other countries, early testing strategies focused only on severe cases or contacts of known cases and those with known international travel, leading to positively biased CFR estimates. A high CFR of 7.2% reported in Italy was attributed to a greater proportion of the elderly in the population and a stringent testing strategy restricted to severe disease cases^4^. With concurrent outbreaks occurring globally, marked discrepancies in CFR became increasingly apparent. In east Asian countries (Vietnam, Thailand, and Philippines), early rises in case incidence have not been followed by similarly sharp CFR increases. CFR estimation is sensitive to testing strategies, initiation of distancing measures, access to healthcare and population age structure. We surmised that since susceptibility to COVID-19 infection extends to the entire population, crude national COVID-19-specific mortality within the country-specific population would be an informative outcome indicator to study differences in mortality patterns amongst countries, in addition to *a priori* defined potential exposure variables including BCG vaccine use in national immunization schedules.

Among vaccination strategies worldwide, *Bacille Calmette Guérin* (BCG) vaccine has the widest use and is accompanied by a strong safety profile. It is given in infancy for prevention of severe forms of tuberculosis. Epidemiological and randomized trial evidence suggest a protective effect of BCG on infant mortality via nonspecific heterologous protection against other infections^5^ possibly through innate immune epigenetic mechanisms^6,7^. BCG lowers experimental viremia in adult human volunteers through upregulation of interleukins such as IL-1β^8^. In a randomized placebo-controlled trial in Indonesia, BCG given monthly consecutively for 3 months significantly reduced incidence of acute upper respiratory infections among individuals aged >65years^9^. In a trial among Native Americans, BCG vaccination given during childhood showed efficacy in preventing tuberculosis up to 60 years after vaccination, indicating the durability of its protection^10^. Demonstration that exposure to BCG vaccination can ameliorate severe COVID-19 disease and lower mortality could rationalize a therapeutic or preventive strategy that can have immediately deployable global impact. Therefore, using existing publicly available data we examined at the ecological level whether country-level COVID-19 mortality was associated with BCG use in national immunization schedules.

## Methods

National COVID-19-attributable death counts as reported on 29 March 2020 from the Johns Hopkins Coronavirus Resource Center ^11,12^ for the top 50 countries reporting highest case events were used to calculate crude COVID-19-attributable mortality per 1 million population. Population and economic data from 2018 (gross domestic product (GDP), and high, middle or low-income status) were derived from the World Bank Population Data and Open Data repository^13^. In order to mitigate the bias centered around the differential epidemic time curves experienced by the different countries, we calculated days from the 100^th^ COVID-19-positive case to align the countries on a more comparable time curve. We included data on BCG vaccine inclusion in national immunization schedules from the BCG World Atlas^14^.

To evaluate the effect of BCG vaccine on mortality attributable to COVID-19, we built a simple log-linear regression model using crude COVID-19-attributable mortality data per 1 million population for each country as outcome, BCG vaccine inclusion in the national immunization schedule as exposure, and adjusted for the effects of the following variables on mortality: country-specific GDP per capita, the percentage of population 65 years and above, and the relative position of each country on the epidemic timeline (days since 100^th^ case reported as of 29 March 2020). The Shapiro-Wilk normality test confirmed that the log-transformed COVID-19-specific mortality was normally distributed. As China, a BCG-using country with a large population and a relatively concentrated death count in a single province can conceivably demonstrate an artifically lowered mortality and skew these results towards BCG use, we did a sensitivity analysis by running our model with the same covariate adjustments but without China. All data were analysed using the R environment for statistical computing (R Core Team 2018)^15^.

## Results

The median crude COVID-19 mortality per 1 million population among countries with economies classified as low-middle-income, upper-midle-income and high-income countries (LMIC, UMIC, HIC) were 0.4 (Interquartile Range (IQR) 0.06, 0.4), 0.65 (IQR 0.2, 2.2) and 5.5 (IQR 1.6, 13.9), respectively (Fig. 1). Characteristics of BCG using and non-BCG using countries with respect to the variables considered are shown in supplementary Table S1.

**Figure 1:**
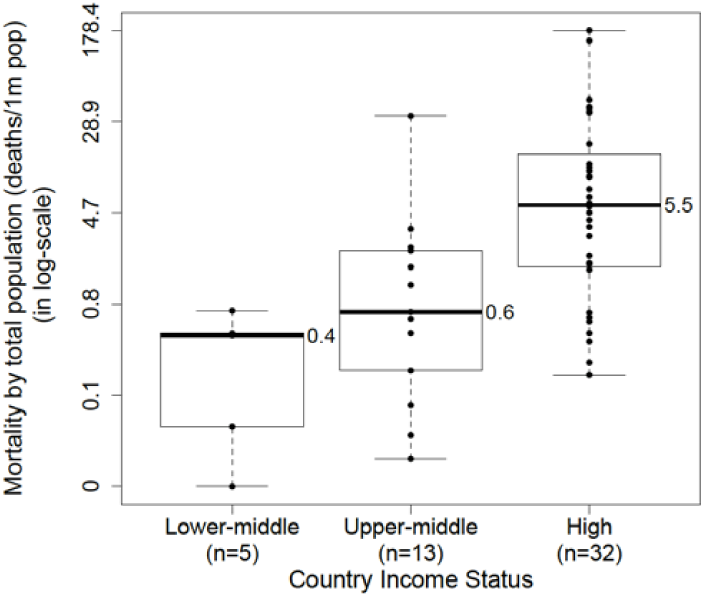
Country economic category and COVID-19-attributable mortality per 1 million population

Plotting mortality per 1 million population by BCG use against percentage of the population ≥ 65 years (Fig. S1) and the relative position of each country in the epidemiological timeline (Fig. S2)) along with the characteristics of countries by BCG status (Table S1) suggested possible confounding effects of these variables. The associations of these potentially confounding variables, the outcome and the exposure of interest are represented visually in Fig. 2.

**Figure 2:**
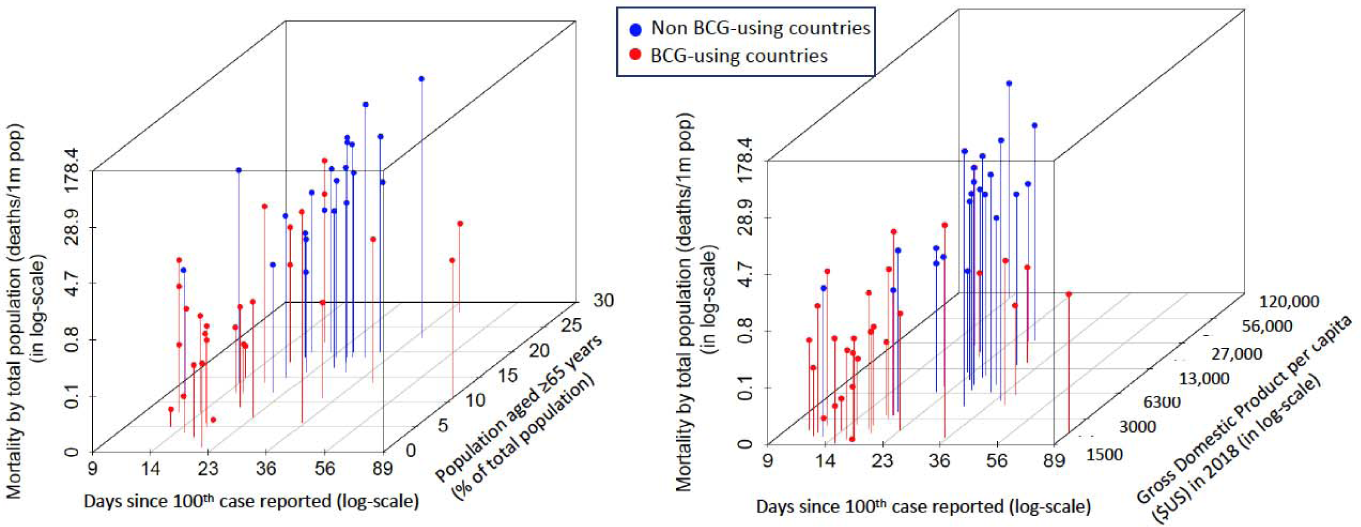
Association between COVID-19-attributable mortality and BCG use in national immunization schedules, propotion of population aged ≥65years, time trajectory on the epidemiological curve and country-specific GDP per capita. Each dot is representative of a country. Red dots=BCG-using countries; Blue dots=Non BCG-using countries.

In the log-linear regression adjusted for per capita GDP, age, and time since 100^th^ case, COVID-19-attributable mortality among BCG-using countries was 5.8 times lower [95% Confidence Intervals (CI) 1.8-19.0; p value = 0.006] than in non BCG-using countries (Table 1). The sensitivity analysis run when excluding China from the model resulted in no appreciable change in the BCG effect (5.7 [95% CI 1.8-18.5]).

**Table 1:** Log-linear regression model of COVID-19-attributable mortality per 1 million population and BCG use in national immunization schedules, adjusting for relevant covariates. Exponentiated estimates for all parameters are provided.

| **Covariate** | **Estimate** | **95% CI** |
| --- | --- | --- |
| BCG non-use in NIP (Reference group was BCG using countries) | 5.78 | 1.75, 19.02 |
| GDP per capita | 1.00 | 0.99, 1.00 |
| Percentage of those aged 65 years and above among total population | 1.08 | 0.99, 1.17 |
| Days since 100<sup>th</sup> reported case | 1.04 | 1.00, 1.08 |

BCG = Bacille Calmette Guérin vaccine; NIP = national immunization program; GDP = Gross Domestic Product; CI = Confidence Interval

## Discussion

The direct association between COVID-19-attributable mortality and country-level economic status observed in this analysis is starkly counterintuitive. Prior global disease burden assessments have suggested that deaths from acute respiratory illness are typically higher in low-income settings due to multiple socio-demographic and economic risk factors^16,17^. Among observed COVID-19-attributable risk factors for disease severity and death, age over 65 years has been identified as a significant factor ^4,18^, while it is inferred that LMICs which typically have a younger population structure would potentially experience fewer overall deaths. Another potential confounder was the time lag in deaths following detection of cases. We selected countries with at least 100 reported cases and adjusted for time since this sentinel event. After adjusting for country economic status, proportion of older population and aligning the epidemic trajectories of the highest hit countries, the intriguing observation of a significant association between BCG use and lower COVID-19-attributable mortality remained discernable.

Recent non-peer reviewed work reported a similar negative association between BCG use and COVID-19 mortality^19^. Unlike our report, their findings did not account for potential confounding effects of income status, age structure of the population or timing of the epidemic, and included only 5 BCG non-using countries, making reliable inference challenging. Currently, BCG vaccine is being considered for clinical trials in different settings to test its ability to mitigate infection effects or protect healthcare workers and the elderly population against the SARS-CoV-2 disease^20^.

Several other determinants related to host, viral and environmental factors may be ascribed to these mortality differences. Prevalence of comorbidities such as diabetes, cardiovascular disease, chronic respiratory disease and cancer are rising in LMICs, and in some settings have overtaken HIC. In 2019 non-insulin dependant diabetes mellitus prevalence among adults was reported to be 11% in Asian countries and 6% in Europe^21^. The World Health Organization estimates higher cardiovascular disease risk in Asian regions than in European and North American regions^22^. Since we had no access to comprehensive nationally representative comorbidity data, the contribution of disease comorbidity as a population level risk factor for COVID-19-attributable mortality remains unclear.

Genetic risk factors associated with susceptibility to SARS-CoV-2 (in CCL2, mannose binding lectin, CXCL10/IP-10 or ACE2 receptor) are currently under evaluation, and as more evidence accumulates, the role played by these factors may become evident^23,24^. Variations in SARS-CoV2 receptor binding domain of the spike protein or nucleocapsid protein could alter disease severity^25^,^26^, although this is more likely to occur in an endemic setting rather than during a pandemic. Temperature and relative humidity may be inversely associated with viral transmissibility. but the association has been modest and inconsistent^27,28^.

Early disease models that used assumptions based on CFRs from China and Italy predicted higher mortality in LMICs, which led to countries adopting severe lockdown measures^29^. Ongoing containment measures are critical for infection transmission mitigation. These measures should be balanced against predicted increases in non-COVID-19 mortality arising directly from economic shutdowns and distancing measures. Severe trade restrictions and lowered productivity can increase poverty and food insecurity globally^30^. Major and prolonged disruptions in crucial health service delivery such as immunization programs or access to emergency obstetric and newborn care can result in a direct increase in preventable deaths, as occurred during and after ebolavirus epidemics^31^. Balancing transmission mitigation against sustaining basic health and nutrition access is a difficult but urgent task.

The limitations in our analyses are important to consider. Deaths lag behind symptomatic infection by 2-8 weeks, and when compared with concurrent incidence cases may underestimate the CFR, although this is less likely to influence cumulative crude mortality^32^. Health system preparedness of each country and the institution of control measures such as social distancing and lockdowns can also determine the cases and mortality numbers. Our data are not meant to falsely reassure countries that their use of BCG may lead to lower mortality. Indeed, our analysis is ecological, does not take into account present BCG coverage, nor timing of BCG vaccine introduction into national schedules, and is not based on a randomized comparison. By far the most important source of unmeasured confounding in our analysis relates to differential testing and reporting. Limited laboratory surveillance availability and access to facility-based care is common in countries using BCG. Substantial case underascertainment or under-reporting of deaths can magnify any association between mortality and BCG use. In exponential functions, small iterations in time result in substantial changes in outcome. Our findings need to be interpreted with caution; given vulnerable health systems and high levels of comorbidities in LMICs, if an exponential rise of cases followed by deaths were to occur in ensuing weeks, this would alter the epidemiological predictions in this report.

Despite all these caveats, the inverse relationship between country economic status and COVID-19-attributable mortality, and the strong ecological association with BCG vaccination are intriguing. The findings warrant deeper epidemiological scrutiny and prospective evaluation in individually randomized trials. Importantly the findings in this report illustrate the pivotal role that continuous systematic laboratory surveillance will have in improving our understanding of the pandemic, particularly in LMICs. Such data lead to informed policy making that are beneficial to health and economic outcomes.

## Data Availability

All data used in this report are based on publicly available data. Corresponding citations for where the data can be accessed are given in the manuscript.

https://coronavirus.jhu.edu/map.html

https://data.worldbank.org/indicator/SP.POP.TOTL?most_recent_value_desc=true

http://www.bcgatlas.org/

